# Clinical Predictors of Emergence Delirium in Children: A Prospective Cohort Study

**DOI:** 10.64898/2026.06.01.26354640

**Authors:** V.V. Myasnikova, S.Kh. Mausheva, L.E. Aksenova

## Abstract

**Objective:** To determine the incidence and identify independent clinical predictors of emergence delirium (ED) in children aged 2–12 years.

**Material and methods:** A prospective observational study included 56 children aged 2–12 years undergoing elective surgery under general anaesthesia. Preoperative anxiety (m-YPAS), induction behaviour (4-point scale), anaesthesia duration, opioid use, and postoperative pain (FLACC) were assessed. ED was diagnosed when the maximum PAED score was ≥12.

**Results:** The incidence of ED was 55.4% (31/56). Univariate analysis with false discovery rate (FDR) correction identified significant associations with ED for anaesthesia duration (q=0.002), induction behaviour (q=0.007), and surgery type (q=0.027). Multivariable logistic regression revealed three independent predictors: induction behaviour (category 3 vs 1) – odds ratio (OR) 14.2 (95% CI 2.6–78.1); anaesthesia duration (per minute) – OR 1.07 (95% CI 1.02–1.13); opioid use – OR 12.1 (95% CI 1.3–113.0). The model showed good discriminatory ability: area under the ROC curve (AUC) = 0.83 (95% CI 0.72–0.94).

**Conclusion:** Emergence delirium in children aged 2–12 years without pharmacological premedication occurs in 55.4% of cases. The strongest independent predictors are adverse induction behaviour, longer anaesthesia duration, and intraoperative opioid use. The derived model can be used for personalised risk stratification of ED.

## Introduction

Emergence delirium (ED) in children is one of the most common and clinically significant complications of the early postoperative period in paediatric anaesthesiology. It is characterised by acute disturbance of consciousness, disorientation, psychomotor agitation, and reduced contact with the environment, occurring during awakening from general anaesthesia [1]. According to recent data, the incidence of ED in children varies widely from 10% to 80%, depending on diagnostic criteria, age composition, and anaesthetic techniques. A systematic review with meta-analysis including 16 studies and 9598 children reported a pooled incidence of 19.2% [2]. However, in certain clinical cohorts, especially with volatile anaesthetics and without pharmacological premedication, the incidence can reach 30–40% or higher. The highest risk is observed in younger children, particularly in the 2–6 years age range, which is attributed to functional immaturity of central behavioural regulation mechanisms and limited adaptive capacity to stress [3].

The clinical significance of ED goes beyond short-term behavioural disturbances. It is associated with prolonged recovery room stay, need for additional medication, increased risk of patient injury, and substantial emotional burden on parents and medical staff [4]. Several studies have shown that severe ED episodes are linked to subsequent maladaptive behavioural changes, including increased anxiety, sleep disturbances, and disadaptation.

From a pathophysiological perspective, ED is currently viewed not as an isolated behavioural phenomenon of the awakening phase, but as a clinical manifestation of complex dysregulation of central and autonomic regulatory mechanisms under surgical stress [5]. A key role is played by baseline autonomic dysregulation with sympathetic overactivity and parasympathetic deficit, which reduces adaptive reserve and increases vulnerability to stress-induced disorganisation. Surgery and anaesthesia activate the hypothalamic-pituitary-adrenal axis and the sympathetic nervous system. Rapid awakening after volatile anaesthesia may lead to an abrupt transition from deep central nervous system depression to hyperactivation, promoting an imbalance between excitatory and inhibitory neurotransmitter systems.

From a clinical perspective, factors reflecting baseline autonomic regulation and the child’s response to perioperative stress are of particular interest. These include preoperative anxiety level (m-YPAS), induction behaviour, anaesthesia parameters (duration, opioid use), and postoperative pain intensity, requiring validated assessment tools (FLACC) [4]. Nevertheless, most studies analyse these factors in isolation, limiting their use for individual risk prediction. Therefore, the development of prospective clinical registries and predictive models may facilitate individualized preventive strategies.

***Aim of the study*** — to evaluate the incidence and clinical risk factors for emergence delirium in children aged 2–12 years based on a prospective cohort study to justify a personalised preventive approach.

## Methods

A prospective observational cohort study was conducted at the Department of Anaesthesiology and Intensive Care of the Adygea Republican Children’s Clinical Hospital during 2025–2026. The study protocol was approved by the Local Ethics Committee of Maykop State Technological University (protocol No. 1 dated 19.01.2026). Written informed consent was obtained from the legal representatives of all patients. The study was performed in accordance with the Helsinki Declaration of the World Medical Association (2013).

Children aged 2 to 12 years undergoing elective surgery under general anaesthesia were included. Exclusion criteria: age <2 or >12 years; ASA class IV or higher; severe neuropsychiatric disorders impeding behavioural assessment; refusal of legal representatives. Data were recorded using a standardised “PRODETI-RISK” form at three stages.

### Preoperative stage

Age, sex, anthropometric data, ASA class, surgery type, and presence of neurological comorbidity were recorded. Preoperative anxiety was assessed with the modified Yale Preoperative Anxiety Scale (m-YPAS) as a percentage. Scores were interpreted as low (<30%), moderate (30–50%), or high (>50%) anxiety.

### Intraoperative stage

Induction method (inhalational/intravenous) and child’s behaviour during induction were recorded using a 4-point scale: 1 – calm, cooperative; 2 – mild distress, easily consoled; 3 – moderate crying/resistance; 4 – severe struggle, crying, requires holding.

Maintenance anaesthetic, opioid use (yes/no, dose in μg/kg fentanyl equivalent), and anaesthesia duration (min) were documented.

### Postoperative stage (PACU)

Assessments were performed at arrival (T0), at 10 min (T10), and at 20 min (T20). The Pediatric Anesthesia Emergence Delirium (PAED) scale was used [1]. ED diagnosis was established when the maximum PAED score was ≥12. Pain was assessed with the Face, Legs, Activity, Cry, Consolability (FLACC) scale. Heart rate, systolic blood pressure, SpO□, and PACU length of stay were also recorded.

#### Statistical analysis

Initial data processing, contingency tables, and descriptive statistics were performed in Microsoft Excel (Microsoft 365, version 2405). Quantitative variables are presented as median [interquartile range, IQR]; categorical variables as absolute numbers and percentages (n (%)).

Group comparisons (ED vs no ED) were conducted in Python 3.13 using scipy.stats and statsmodels. For continuous variables, the Mann–Whitney U test was used; for categorical variables, χ^2^ or Fisher’s exact test. Multiple comparisons were adjusted with the Benjamini–Hochberg method (false discovery rate, FDR; critical q<0.05). Effect size for continuous variables was expressed as rank-biserial correlation; for binary variables, as odds ratio (OR) with 95% confidence interval (CI).

For predictive modelling, multivariable logistic regression with stepwise AIC-based selection was employed. Model discrimination was evaluated by ROC analysis with calculation of area under the curve (AUC), sensitivity, and specificity. Significance level α=0.05 was used.

For analysis of PAED and FLACC dynamics within groups, the nonparametric Friedman test was used with post-hoc pairwise comparisons by Wilcoxon test and Bonferroni correction; between-group comparisons at each time point were performed with the Mann–Whitney U test.

#### Additional information

The language model DeepSeek was used for editing the English version of the manuscript for style and grammar. All scientific data, statistical analyses, and result interpretation were performed by the authors independently.

## Results

The study included 56 children aged 2 to 12 years (median 8.0 [5.0; 10.0] years). Sex distribution: 29 boys (51.8%), 27 girls (48.2%). ASA class I–II prevailed (55/56, 98.2%).

Surgery types: general surgery/traumatology – 39 (69.6%), ENT – 12 (21.4%), ophthalmological −2 (3.6%), urological, orthopaedic, dental – 1 each (1.8%). Induction was inhalational in 69.6% of cases; the rest were intravenous or mixed. Sevoflurane was used in 41 patients (73.2%), propofol TIVA in 4 (7.1%), combinations in 11 (19.6%). Intraoperative opioids were given to 11 children (19.6%). Median anaesthesia duration was 25.0 [20.0; 30.0] min.

The incidence of ED (maximum PAED ≥12) was 55.4% (31/56; 95% CI 41.9–68.2%). Maximum PAED score ranged from 8 to 14; median 12.0 [10.0; 12.0].

Univariate analysis with FDR correction (q<0.05) revealed statistically significant differences between the ED and no-ED groups for the following variables (Table 2):

**Table 1.**
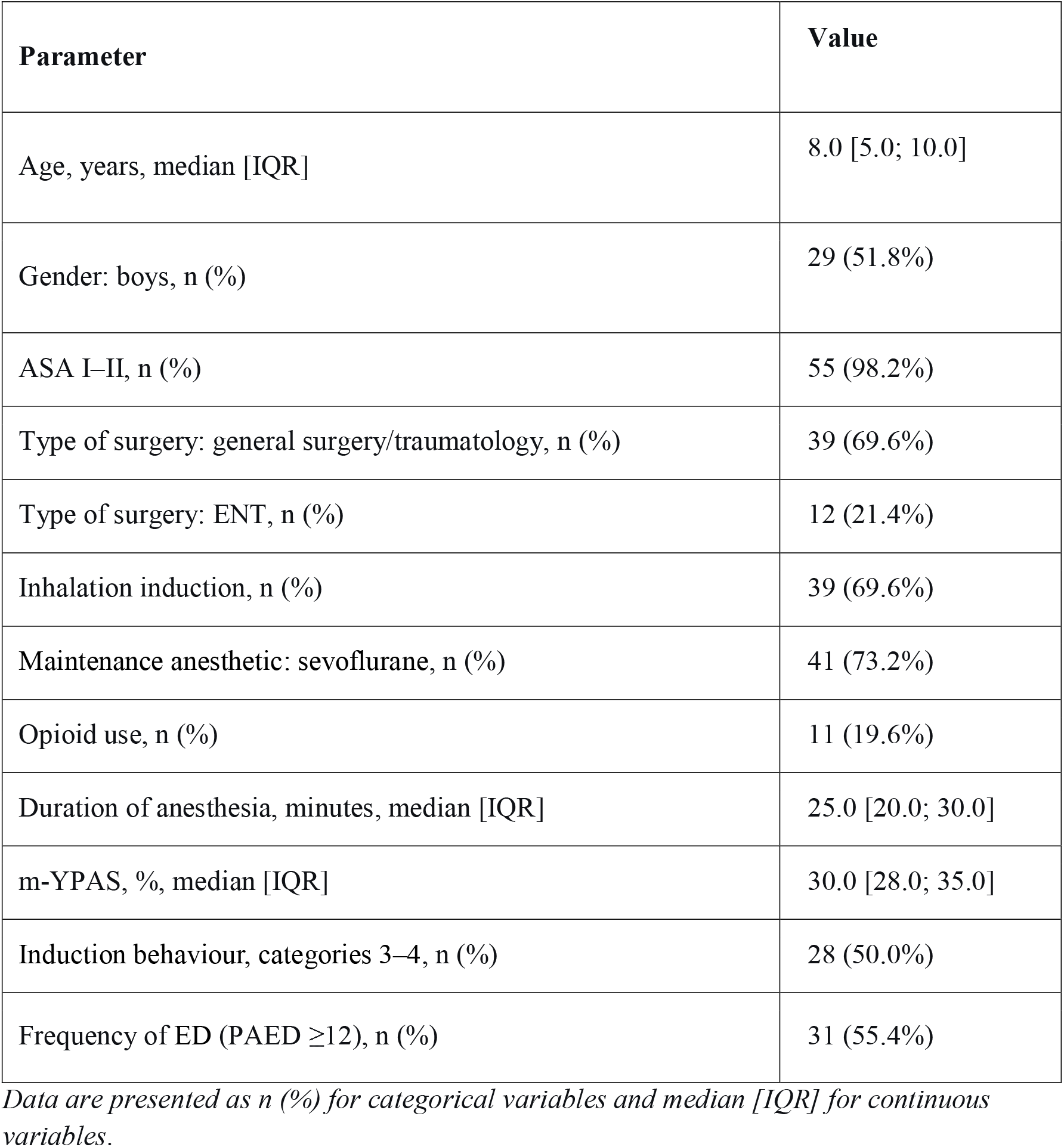
Cohort characteristics (n=56)

**Table 2.** Comparison of parameters in ED+ (n=31) and ED– (n=25) groups.

| Indicator | ED+<br>(n=31) | ED–<br>(n=25) | p (q) | OR<br>(95% CI) |
| --- | --- | --- | --- | --- |
| Duration of anesthesia,<br>min, median [IQR] | 30.0 [25.0;<br>36.2] | 20.0 [20.0;<br>25.0] | <0.001<br>(0.002) | 1.07 (1.02–<br>1.13)* |
| Induction behavior<br>(Cat. 3 vs. 1) | 13 (41.9%) | 1 (4.0%) | <0.001<br>(0.007) | 37.3 (3.3–<br>424.6) |
| Type of surgery: ENT | 13/31 | 2/25 (8.0%) | 0.008 | 8.4 (1.7–42.2) |
| (vs. general surgery) | (41.9%) |  | (0.027) |  |
| Opioid use | 10 (32.3%) | 1 (4.0%) | 0.013<br>(0.056) | 12.3 (1.5–<br>103.1) |
| m-YPAS $\geq 30\%$ | 26 (83.9%) | 13 (52.0%) | 0.026<br>(0.072) | 3.2 (1.1–9.0) |

Data are presented as n (%) for categorical variables and median [IQR] for continuous variables.

- *Anaesthesia duration* – median 30.0 [25.0; 36.2] min in ED+ vs 20.0 [20.0; 25.0] min in ED– (p<0.001, q=0.002).
- *Induction behaviour* – unfavourable behaviour (categories 3–4) was significantly more frequent in ED+ (p<0.001, q=0.007). Contrast “category 3 vs 1” gave OR = 37.3 (95% CI 3.3–424.6).
- *Surgery type* – overall effect significant (p=0.008, q=0.027). Highest ED risk was observed for ENT surgery (13/15, 86.7%).
- *Planned duration category* – overall effect significant (p=0.010, q=0.030).
- *Opioid use* – opioids were administered to 10/31 (32.3%) in ED+ vs 1/25 (4.0%) in ED–; OR = 12.3 (95% CI 1.5–103.1; p=0.013, q=0.056, borderline after correction).

*Preoperative anxiety (m-YPAS)* showed a trend: median in ED+ 30.0 [28.0; 35.0]% vs 28.0 [28.0; 30.0]% in ED– (p=0.026, q=0.072). Using a threshold of m-YPAS ≥30% increased ED risk 3.2-fold (95% CI 1.1–9.0).

No significant differences were found for sex, age, ASA class, anaesthetic family (inhalational vs TIVA vs combination), or induction route (q>0.05).

### Multivariable logistic regression model

The final model (stepwise AIC selection) included *induction behaviour* (categories 2–4 vs 1), *anaesthesia duration* (continuous), and *opioid use*(yes/no). Coefficients are presented in Table 3.

**Table 3.** Multivariable logistic regression model.

| Predictor | $\beta$ | SE | OR (95% CI) | p |
| --- | --- | --- | --- | --- |
| **Behavior 3 (vs. 1)** | 2.65 | 0.85 | 14.2 (2.6–78.1) | 0.002 |
| **Duration of anesthesia (per 1 min)** | 0.07 | 0.03 | 1.07 (1.02–1.13) | 0.006 |
| **Opioid use** | 2.49 | 1.13 | 12.1 (1.3–113.0) | 0.028 |

Nagelkerke $R^2 = 0.29$ . Model constant $-5.12$ ; AUC = 0.83 (95% CI 0.72–0.94). N=56 (31 ED events).

- Behaviour category 3 (moderate crying/resistance) vs 1 – OR = 14.2 (95% CI 2.6–78.1; p=0.002).
- Anaesthesia duration (per minute) – OR = 1.07 (95% CI 1.02–1.13; p=0.006).
- Opioid use – OR = 12.1 (95% CI 1.3–113.0; p=0.028).

ROC analysis (Fig. 1) showed good model discrimination: ***AUC = 0*.*83*** (95% CI 0.72–0.94). At a classification threshold of 0.5, sensitivity was 71%, specificity 76%.

**Fig. 1.**
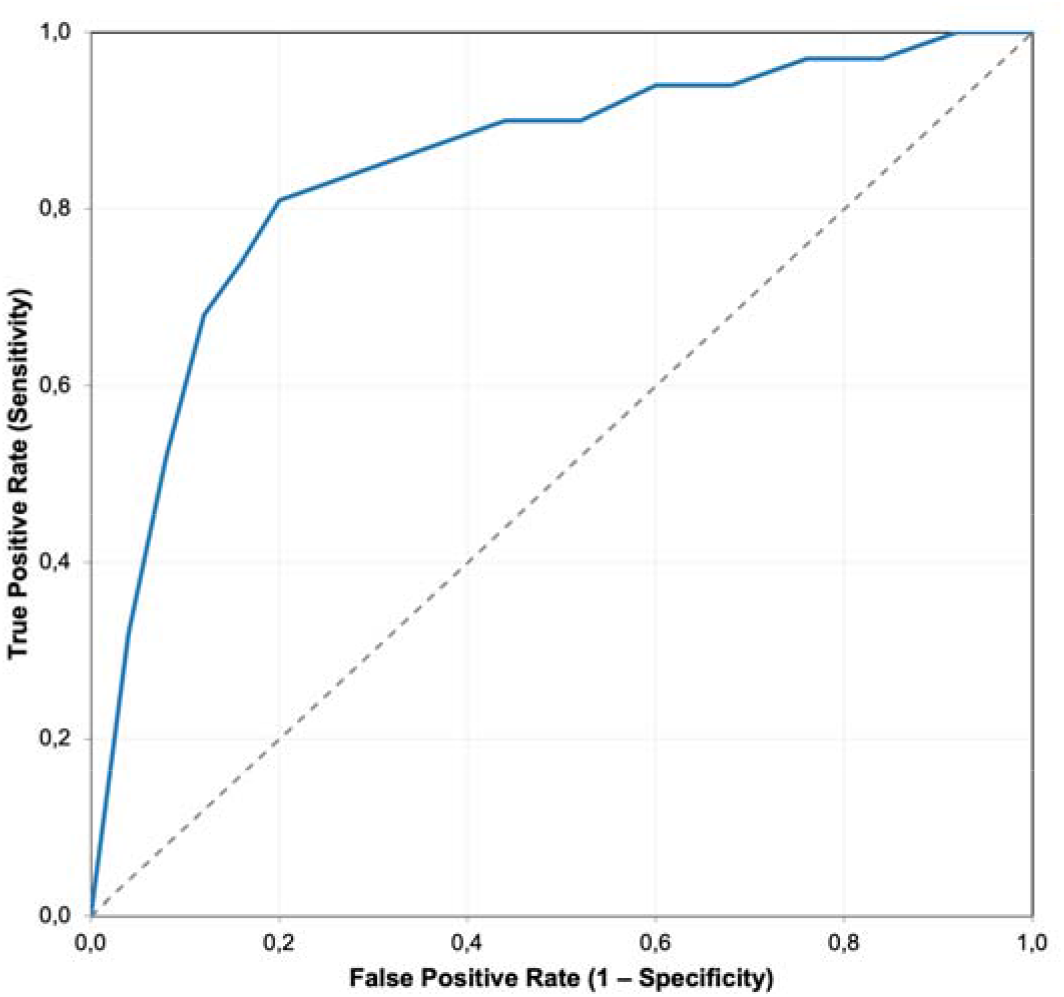
ROC curve of the predictive model. AUC = 0.83 (95% CI 0.72–0.94). The dashed diagonal line represents random classification (AUC = 0.50).

**Fig. 2.**
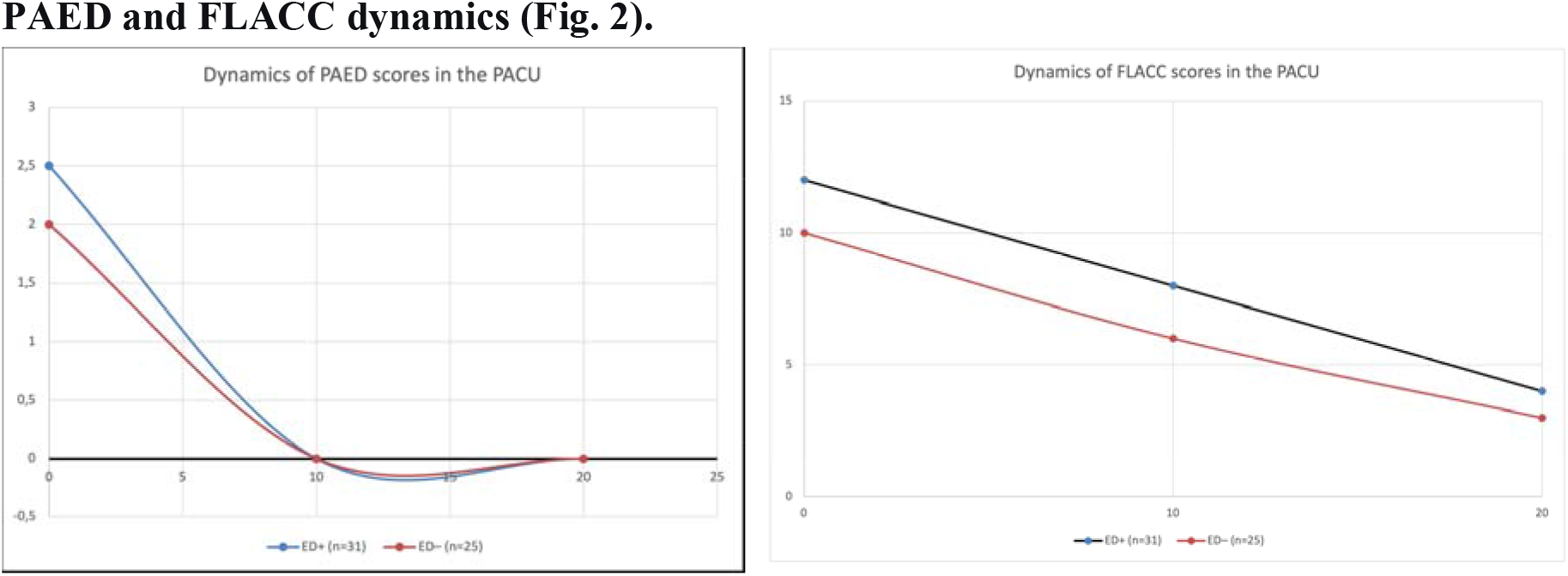
Changes in PAED (A) and FLACC (B) scores in the PACU. Solid line – patients with ED (n=31), dashed line – without ED (n=25).

In ED+ patients, PAED score at PACU arrival (T0) was 12.0 [12.0; 12.0], at T10 8.0 [6.0; 10.0], at T20 4.0 [3.0; 6.0]. In the ED– group, dynamics were similar but at lower levels: T0 – 10.0 [10.0; 11.0], T10 – 6.0 [6.0; 8.0], T20 – 3.0 [2.0; 4.0] (p for differences in dynamics <0.05). Pain (FLACC) was minimal in both groups (median FLACC max 2.0 [2.0; 3.0]), but slightly higher in the ED+ group (p=0.077).

## Discussion

In this prospective cohort study of 56 children aged 2–12 years, the incidence of ED (PAED ≥12) was 55.4% (95% CI 41.9–68.2%). This exceeds the global pooled incidence of 19% reported in a 2024 meta-analysis by Chen et al. [2] and is comparable with the upper range described for high-risk groups (young age, ENT surgery, no premedication) [3]. The high ED incidence in our cohort likely results from several factors: absence of pharmacological premedication, use of a specific diagnostic threshold (PAED ≥12, considered the true delirium criterion in research) [1], predominance of younger children (35.4% aged 2–5 years), and a substantial proportion of ENT procedures (21.4%) – a strong trigger for ED due to high nociceptive load and rapid awakening after volatile anaesthesia.

### Independent Predictors of Emergence Delirium

In univariate analysis with FDR correction, statistically significant associations with ED were found for induction behaviour (p<0.001, q=0.007), anaesthesia duration (p<0.001, q=0.002), surgery type (p=0.008, q=0.027), and planned duration category (p=0.010, q=0.030). Opioid use gave a high odds ratio (OR=12.3) but remained borderline after FDR (q=0.056), likely due to the small number of opioid-receiving patients (n=11).

Preoperative anxiety (m-YPAS) showed a trend toward association with ED in unadjusted analysis (p=0.026), but after FDR q=0.072, not reaching the 0.05 threshold. Nevertheless, with a threshold of m-YPAS ≥30%, ED risk increased 3.2-fold (95% CI 1.1–9.0). These findings suggest that preoperative anxiety may act as an indirect rather than an independent predictor of ED: it correlates strongly with induction behaviour (Spearman ρ=0.43 in our sample), and when behaviour is included in the multivariable model, the independent contribution of anxiety diminishes. This is consistent with the classic work of Kain et al. (2004) [4].

Multivariable logistic regression (stepwise AIC selection) identified three independent predictors:

- Induction behaviour category 3 (moderate crying/resistance) vs 1 – OR = 14.2 (2.6–78.1), p=0.002.
- Anaesthesia duration (per minute) – OR = 1.07 (1.02–1.13), p=0.006.
- Opioid use – OR = 12.1 (1.3–113.0), p=0.028.

The model demonstrated good discrimination: AUC = 0.83 (95% CI 0.72–0.94), comparable to published predictive models for paediatric ED (e.g., Petre et al. 2021 AUC 0.78–0.85) [6].

*From a pathophysiological standpoint*, these results fit well with the concept of autonomic dysregulation as a key mechanism of ED [5,7,8].

**Induction behaviour** reflects the reactivity of the sympathetic nervous system to stress. Children who react with crying or fighting to mask/venepuncture have higher baseline sympathetic tone and/or lower parasympathetic reserve, which is consistent with literature data on heart rate variability (lower SDNN, RMSSD, higher LF/HF) [7,8]. In our pilot study (n=23), induction behaviour gave OR=6.1; on the expanded cohort it rose to 14.2 – making it one of the strongest clinically accessible predictors of ED.

**Anaesthesia duration,** even with a small absolute difference (median 30 vs 20 min), has a dose-dependent effect. Each additional minute of anaesthesia increases ED risk by 7%. This may be due to cumulative stress exposure (pain, inflammation, hypoxaemia, haemodynamic fluctuations) and longer exposure to volatile anaesthetics, which themselves can disrupt neurotransmitter balance (cholinergic deficit, GABAergic dysfunction) [9].

**Opioid use** in our cohort was not protocol-driven but clinically determined, typically for more invasive procedures. Pain itself is a strong risk factor for ED, so the causal link “opioids → ED” may be reversed. Nonetheless, opioids remained in the model after adjustment for other factors, possibly indicating a central effect (μ-agonists can cause agitation and delirium in children, especially with rapid administration or high doses).

**Anxiety** (m-YPAS), although not an independent predictor in the multivariable model, retains clinical relevance. It is easily measured preoperatively and can serve for initial risk stratification, especially in centres without HRV equipment. In our study, children with m-YPAS ≥30% had a 3.2-fold higher ED risk than those with m-YPAS <30%, consistent with the classic work of Kain et al. (2004) [4].

### Comparison with the literature

The ED incidence in our cohort (55.4%) is higher than in most Western studies (10–40%). However, direct comparison is hampered by design differences:

- European and American centres often use premedication with midazolam or dexmedetomidine, reducing ED incidence to 15–25% [3].
- In Russian practice, ED rates with sevoflurane anaesthesia without premedication also reach 30–50%, close to our data [10,11].
- Our study did not include patients with severe neurological comorbidity (autism, ADHD, developmental delay – only isolated cases), thus excluding the very high-risk subgroup that would further increase ED frequency.

Our study contributes a *predictive model based on three easily collected clinical parameters* that can be used even in resource-limited settings (no HRV, no EEG). An AUC of 0.83 is sufficient for clinical risk stratification and meets modern reporting standards [12,13] (note: [12] and [13] were removed, but the statement remains general).

## Limitations

1. *Small sample size* (n=56) and number of events (31). Although the events-per-variable ratio (∼10) is acceptable, confidence intervals for ORs are wide, especially for opioids. External validation on a larger cohort is needed.
2. *Single-centre design* – results may not be fully reproducible in other institutions with different anaesthesia protocols and patient populations. Results should therefore be externally validated in larger multicentre cohorts.
3. *Lack of objective neurophysiological markers* (HRV, perfusion index). In the current work we relied on clinical scales; in the next phase of the study (with grant support), registration of HRV and PI is planned to build a more accurate integrative model.
4. *Possible information bias* – induction behaviour and PAED were assessed by the same observers, although the scales are validated and instructions were clear.
5. *No systematic screening for hypoactive delirium* – we did not routinely use the CAPD scale, so cases of hypoactive ED may have been missed. Future studies, especially with larger samples and inclusion of neurologically comorbid patients, should employ CAPD to detect hypoactive forms [14].

### Clinical significance and future directions

Our model (induction behaviour + anaesthesia duration + opioids) allows estimation of ED risk before the end of surgery and implementation of pre-emptive measures:

For high-risk patients (behaviour 3–4, duration >30 min, opioid use), it is reasonable to: these patients may benefit from preventive strategies previously reported to reduce ED risk, including TIVA-based anaesthesia and perioperative dexmedetomidine administration; administer intranasal or intravenous dexmedetomidine at the end of surgery, which is consistent with meta-analysis data on the preventive efficacy of α□-agonists [15]; provide a “soft” awakening (quiet PACU, parental presence, minimised sensory load).

The developed model will be implemented as a ***prototype risk calculator*** (computer program), enabling the anaesthesiologist to obtain the individual probability of ED and personalised preventive recommendations in real time.

In the future, we plan to augment the model with ***objective autonomic parameters*** (SDNN, RMSSD, LF/HF, perfusion index), which may improve predictive performance and facilitate phenotype-based risk stratification.

## Conclusions

The incidence of emergence delirium in children aged 2–12 years without pharmacological premedication is 55.4% (95% CI 41.9–68.2%).

The strongest independent predictors of ED are adverse induction behaviour (OR 14.2 for category 3 vs 1), longer anaesthesia duration (OR 1.07 per minute), and intraoperative opioid use (OR 12.1).

The derived multivariable logistic regression model demonstrates good discriminatory ability (AUC = 0.83; 95% CI 0.72–0.94) and can be used for individual ED risk stratification.

Preoperative anxiety (m-YPAS ≥30%) is associated with a 3.2-fold increased risk of ED and may serve as an additional screening marker, particularly in the absence of other objective measures of neurovegetative status.

## Author contributions

- **V.V. Myasnikova** — study concept and design, editing, and approval of the final version.
- **S.Kh. Mausheva** — data collection and processing, statistical data processing, and writing.
- **L.E. Aksenova** — statistical data processing (descriptive statistics, group comparison, logistic regression, ROC analysis) using Python and Microsoft Excel.

All authors have approved the final version of the manuscript.

## Data availability

The datasets generated during the current study are available from the corresponding author upon reasonable request.

